# HumanELY: Human evaluation of LLM yield, using a novel web-based evaluation tool

**DOI:** 10.1101/2023.12.22.23300458

**Authors:** Raghav Awasthi, Shreya Mishra, Dwarikanath Mahapatra, Ashish Khanna, Kamal Maheshwari, Jacek Cywinski, Frank Papay, Piyush Mathur

## Abstract

Large language models (LLMs) have caught the imagination of researchers,developers and public in general the world over with their potential for transformation. Vast amounts of research and development resources are being provided to implement these models in all facets of life. Trained using billions of parameters, various measures of their accuracy and performance have been proposed and used in recent times. While many of the automated natural language assessment parameters measure LLM output performance for use of language, contextual outputs are still hard to measure and quantify. Hence, human evaluation is still an important measure of LLM performance,even though it has been applied variably and inconsistently due to lack of guidance and resource limitations.

To provide a structured way to perform comprehensive human evaluation of LLM output, we propose the first guidance and tool called HumanELY. Our approach and tool built using prior knowledge helps perform evaluation of LLM outputs in a comprehensive, consistent, measurable and comparable manner. HumanELY comprises of five key evaluation metrics: relevance, coverage, coherence, harm and comparison. Additional submetrics within these five key metrics provide for Likert scale based human evaluation of LLM outputs. Our related webtool uses this HumanELY guidance to enable LLM evaluation and provide data for comparison against different users performing human evaluation. While all metrics may not be relevant and pertinent to all outputs, it is important to assess and address their use.

Lastly, we demonstrate comparison of metrics used in HumanELY against some of the recent publications in the healthcare domain. We focused on the healthcare domain due to the need to demonstrate highest levels of accuracy and lowest levels of harm in a comprehensive manner. We anticipate our guidance and tool to be used for any domain where LLMs find an use case.

**Link to the HumanELY Tool:** https://www.brainxai.com/humanely

## 1 Introduction

With the release of ChatGPT in November 2022, the number of similar foundation models and large language models have increased exponentially. Foundation models (FM) are large-scale neural networks trained on broad data (Koubaa et al. [2023]). They may handle multiple types of data (text, image, audio), making them a superset of Large Language Models (LLMs) (Bommasani et al. [2021]). These models which are also accelerating research and implementation at exponential pace are complex with varying levels of output. Although commonly expected to perform better as the number of parameters increase, we are also seeing varying levels of performance as demonstrated on various leaderboards (Ope). The performance of these models varies not only based on the training data, but also on their architecture and specific task. Many use cases and pilots to evaluate performance of these models are being undertaken by researchers from across the world. Numerous evaluation metrics have been used for these models, which regularly leads to new winners being declared (Ope).

Evaluation of LLMs is a challenging task with many proposed technical metrics and human evaluation methods (Kamalloo et al. [2023]) ^1^. As these models continue to evolve, their accurate evaluation is challenging. Most of the models have been assessed using technical methods, not universally designed to evaluate their performance, or do a narrowly performance assessment on a few tasks or domains. There is also growing evidence that many models struggle with robustness and potential for significant harm due to hallucatinations(Guo et al. [2023], Rawte et al. [2023]) ^2^.

In a recent comprehensive survey of LLMs categorized in three major groups, knowledge and capability evaluation,alignment evaluation and safety evaluation, the researchers assessed various aspects of model performance and their applications in certain key areas such as healthcare,finance,legal, etc (Guo et al. [2023]). The authors emphasize that as these LLMs rapidly advance in their capabilities, existing methods for evaluation are still lacking in their holistic assessments. They propose future directions for improvement in model development and evaluation to align with human values of helpfulness,harmlessness and honesty.

While challenging to perform, the current gold standard for evaluation of these models beyond a few technical metrics, is still human evaluation. Human evaluation provides for a comprehensive contextual benchmark beyond the task, domain and semantic capabilities oriented current technical approaches. Human evaluation also aligns with the human values that many of these LLMs have been designed to replicate in their performance. But unlike technical metrics which have a standard design, human evaluations have been designed and performed with significant variations. Multiple human evaluation methods have been proposed and used to assess NLP models, but there is no systematic method specifically for evaluating LLMs (van der Lee et al. [2021], Shimorina and Belz [2021], Fuentes et al. [2022], Bojic et al. [2023]). Additionally, these evaluation tools suffer from critical issues such as reproducibility and generalizability across multiple tasks and domains, including healthcare, finance, education, etc (Bhatt et al. [2021], Ito et al. [2023], Mahamood [2023], Li et al. [2023]).Hence,having a comprehensive,consistent,contextual,efficient and effective design of human evaluation is needed.

To address these challenges, we introduce HumanELY (Human Evaluation of LLM Yield), aiming to provide not only guidance for human evaluation of LLMs but also a state-of-the-art tool to facilitate these evaluations. We offer an exhaustive set of questions with clear definitions and details for each metric in the context of LLM evaluation. Through these efforts, we aim to establish a better platform for human evaluation, enhancing reproducibility and generalizability in the evaluation process.’

### Review of existing evaluation metrics for LLMs

Assessing LLMs is a challenging task, as there is no one-size-fits-all evaluation method (Zhang et al. [2023], Chan et al. [2023]). Researchers have utilized diverse approaches to evaluate these models, combining both quantitative and human evaluation techniques. Numerous quantitative metrics are developed for specific tasks like Text Classification, Natural Language Inference, Natural Language Generation, Text Summarization, Question Answering, Dialogue generation and Machine Translation (**Table 2**). For example, BLEU scores (Papineni et al. [2002]) are valuable for evaluating tasks like Machine Translation and Question Answering, measuring the similarity between generated and reference text. Similarly, ROUGE scores Lin [2004] are used for text summarization by analyzing n-grams. Metrics like BLEURT (Sellam et al. [2020]) have emerged, incorporating BERT-based representations to assess semantic similarity between generated and reference text, allowing for a more nuanced evaluation.

Metrics used for assessing dialogue quality include Deep-AM-FM (Zhang et al. [2021]), FlowScore (Li et al. [2021]). Deep-AM-FM measures dialog quality with Adequacy Metric (AM) and Fluency Metric (FM), utilizing BERT embeddings and language model probabilities. FlowScore calculates the dialogue quality by modeling dynamic information flow in dialogue history. For tasks like text classification, standard classification metrics like Accuracy, F1 score, Precision, Recall, and AUROC are typically used. However, since LLMs are capable of performing multiple tasks, benchmarking models requires more than task-specific metrics. Benchmarks like SuperGLUE (Sarlin et al. [2020]) have been used in models like GPT 3.0 (Brown et al. [2020a]) and BLOOM (Scao et al. [2022]) (**Table 1**) to showcase model performance. SuperGLUE is a comprehensive evaluation benchmark for assessing natural language understanding models, featuring a range of challenging language tasks to assess language models’ understanding and reasoning capabilities. Its aggregated score offers a holistic measure of a model’s language understanding, making it valuable for comparing LLMs.

**Table 1.** Key LLMs with some of the key Evaluation metrics used in their methods.

| LLM | Architecture | Evaluation Metrics |
| --- | --- | --- |
| PaLM (Chowdhery et al. [2023]) | Decoder | BLEURT, MQM |
| BLOOM (Workshop et al. [2022]) | Decoder | SuperGLUE, sacrebleu, ROUGE |
| GPT 3.0 (Brown et al. [2020b]) | Decoder | Accuracy, Bleu, SuperGLEU |
| Flan -T5 (Chung et al. [2022]) | Encoder-Decoder | Accuracy, Normalised preferred metric |
| GPT 4 <sup>7</sup> | Decoder | Accuracy, Likelihood Estimation |
| T0 (Sanh et al. [2021]) | Encoder-Decoder | Accuracy, rank classification |
| Llama2 (Touvron et al. [2023]) | Decoder | ROUGE, Self-BLEU, Gwet’s ½, TruthfulQA, |
| Falcon (Penedo et al. [2023]) | Encoder-Decoder | Zero-shot evaluation <sup>8</sup> |
| MedPalm2 (Singhal et al. [2023]) | Decoder | Accuracy, Paiwise preference Analysis, Overlap Analysis |
| gatortron (Yang et al. [2022]) | Decoder | Precision, Recall, F1 score, Pearson Correlation, Exact Match |

**Table 2.** Evaluation metric with their strengths and weaknesses.

| Type of Metric | Metric | Explanation | Strengths | Weakness |
| --- | --- | --- | --- | --- |
| Model-free metrics | Perplexity | Perplexity measures the quality of the probability distribution of words in a given corpus by a model. Mathematically, perplexity is the geometric mean of the inverse probabilities of all the words predicted by language models. Lower perplexity represents the better performance of the model. | Easy to compute | Difficult to interpret |
|  | BLEU | The BLEU score is used to evaluate the model's capabilities for machine translation. The BLEU score is calculated by comparing the n-grams of the ground truth machine translation with those of the model-generated machine translation. The BLEU score ranges between 0 and 1, and a higher score represents better model performance. | Frequently used in machine translation tasks. | Inability to understand semantic similarity |
|  | ROUGE | The ROUGE score is commonly used to evaluate summarization tasks by calculating the overlapping n-grams between the model-generated summary and the reference summary. The ROUGE score measures the recall of n-grams in the model-generated summary and ranges from 0 to 1, where a higher score represents better performance. | Frequently used in summarization tasks. | Inability to recognize paraphrasing and contextual meaning |
|  | ACCURACY | Accuracy is a simple measure of correctness, commonly used in classifying word tokens. It is presented as a percentage of correct predictions. | Easy to understand | Limited scope in NLP tasks |

<sup>9</sup>https://learn.microsoft.com/en-us/azure/ai-services/language-service/summarization/custom/how-to/test-evaluate

Table 2: Evaluation metric with their strengths and weaknesses.
|  |  |  |  |  |
| --- | --- | --- | --- | --- |
| Model-based metrics | QuestEval | QuestEval is a fact-oriented, reference-less method to evaluate the summarization quality of language models. QuestEval employs a diverse set of questions to assess the correctness and appropriateness of the generated text. | Do not require the ground truth or reference text | Can be biased |
|  | Med-HALT | Med-HALT is specifically designed for the medical domain and calculates the hallucination generated by large language models. | Useful for the medical domain, as the model is domain-specific | Not a method but provides a new hallucination benchmark and dataset |
|  | BERTSCORE | BERTSCORE uses the contextual embeddings generated by the BERT model at the token level for both the model output and reference text and then calculates the cosine similarity between them. Based on the cosine similarity, BERTSCORE computes the Recall, Precision, and F1 score. | Considers the contextual information through embeddings and can be used for any downstream NLP task | Computationally expensive because it uses embeddings |

The Falcon model evaluation strategy (Penedo et al. [2023]) is inspired by The BigScience Architecture & Scaling Group (Scao et al. [2022]) and has employed the unified framework LM-Eval^3^ developed by *EleutherAI*. This framework assesses the large language model across more than 200 tasks and supports models from Hugging Face Transformers^4^, including MegaTron-DeepSpeed^5^ and GPT-NeoX^6^. Hence, LM-Eval provides users with multi-environment flexibility.

While these metrics offer valuable insights into model performance, they may not fully capture the complexities of text, including issues like hallucinations, biases, and ethical concerns. Recognizing these complexities, human evaluation is increasingly seen as a complementary solution. Recent models like Llama2 (Touvron et al. [2023]) and MedPalm (Singhal et al. [2022]) combine human evaluation with quantitative assessment to present a more realistic and comprehensive evaluation of model performance. Additionally, evaluation benchmarks such as ChatbotArena and MT-Bench (Zheng et al. [2023]), proposed for evaluating LLMs and chatbots across multiple tasks, provide task-centric assessments. Chatbot Arena allows users to interact with anonymous LLMs and vote based on their preferences. Whereas, MT-Bench evaluates LLMs on multi-turn dialogues using a comprehensive set of questions tailored to assess their capabilities in handling conversations. Furthermore, a recent evaluation methodology, LLMEval (Lin and Chen [2023]), has introduced a unified approach to evaluation. This approach is based on the creation of a single prompt-based evaluation method that utilizes a unified evaluation framework to encompass various aspects of conversation quality within a single model query. Another noteworthy LLM evaluation metric, HELM, provides a comprehensive assessment of LLMs by evaluating them across various aspects, including language understanding, generation, coherence, context sensitivity, common-sense reasoning, and domain-specific knowledge (Liang et al. [2022]).

Considering the importance of human evaluation, the assessment by experts is still regarded as the gold standard. Human evaluation is subject to variability and inconsistency due to different methods and criteria. This variability can introduce bias in algorithm evaluation and make it challenging to compare different experiments, even though they all fall under the umbrella of “human evaluation”. Various general-purpose human evaluation methods have been proposed, such as Microsoft Human Evaluation ^9^ and LongEval (Krishna et al. [2023]), in their research found that the fine-grained annotations led to lower inter-annotator variance when compared with coarse-grained annotation. Additionally, they also concluded that the partially annotating a summary can reduce annotator workload while maintaining accuracy. They noted that these methods may have limited usefulness for evaluating long-form summaries. Therefore they recommended using reference-free metrics as diagnostic tools for analyzing and understanding model behavior, rather than relying solely on measures of how well models perform specific tasks.

### 1.1 Human Evaluation

Currently, human evaluation still remains the gold standard for the measuring performance of the LLM output,especially in specific domains such as healthcare (Reddy [2023]). Human evaluation can however be variable and inconsistent following different methods and different criteria (Ziems et al. [2023]). Even though many of these methods use the term human evaluation as a homogeneous entity, comparisons between different experiments hard to evaluate and can be biased. Krishna et al., described the numerous challenges with these varying human evaluation methods while evaluating LLM outputs, including inherent subjectivity,inter-rater correlation, labor intensiveness and design challenges. Interestingly, in their comprehensive survey of 162 relevant publications, they found 101 (62%) publications did not perform any human evaluation.They proposed guidelines for human evaluation of faithfulness in long form summarization(150 words or more) (Krishna et al. [2023]). Their key guidelines, termed LongEval, include:

- fine grained annotations have lower inter-annotator variance than COARSE-grained annotations.
- partially annotating a summary reduces annotator workload while maintaining accuracy.
- highlighting hints in the source document has limited usefulness for evaluating long-form summaries.

We use some of this guidance in development of our human evaluation tool, *HumanELY*, to address this and provide a tool for application.

### 2 Proposed Method for consistent and comprehensive human evaluation of LLMs

Considering the importance of human evaluation, in this paper, we propose (**Figure 1**) a structured approach to conducting human evaluation of LLM outputs. We provide a comprehensive list of survey-based questions and majorly divide the human evaluation practice into the following aspects: assessing the relevance of LLM-generated text, evaluating the coverage of LLM-generated text, assessing the coherence of LLM-generated text, considering potential harm associated with LLM-generated text, and emphasizing the significance of comparing human-generated text with LLM-generated text. It is important to understand that different evaluation metrics may overlap; for example, inaccurate answers can be both harmful and unsafe (**Figure 2**).

**Figure 1.**
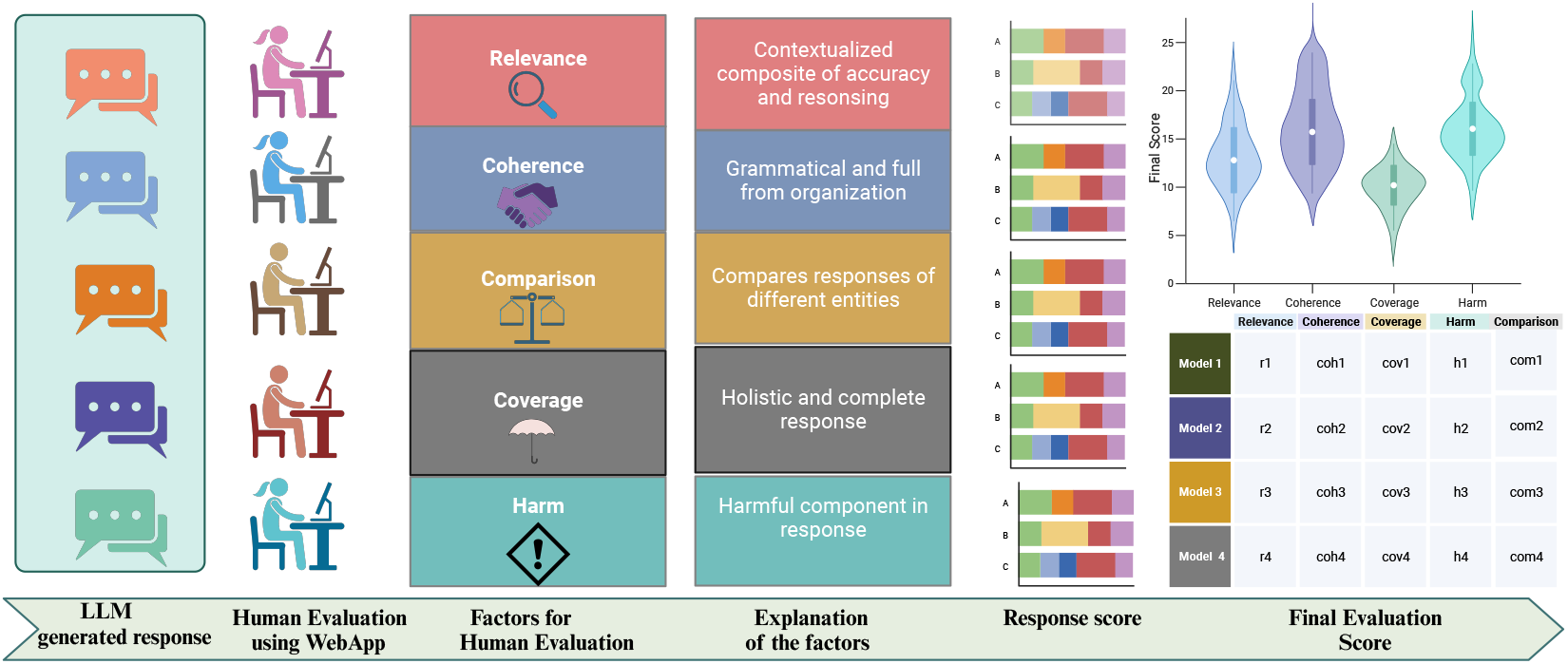
Proposed Method for Human Evaluation Through the HumanELY Portal. We have proposed five major factors for conducting human evaluation: Relevance, Coverage, Coherence, Comparison, and Harm. We have developed a set of survey-based questions to evaluate these five categories. Additionally, we are providing a WebApp that allows for evaluation by simply uploading a file with reference text and human-generated text.

**Figure 2.**
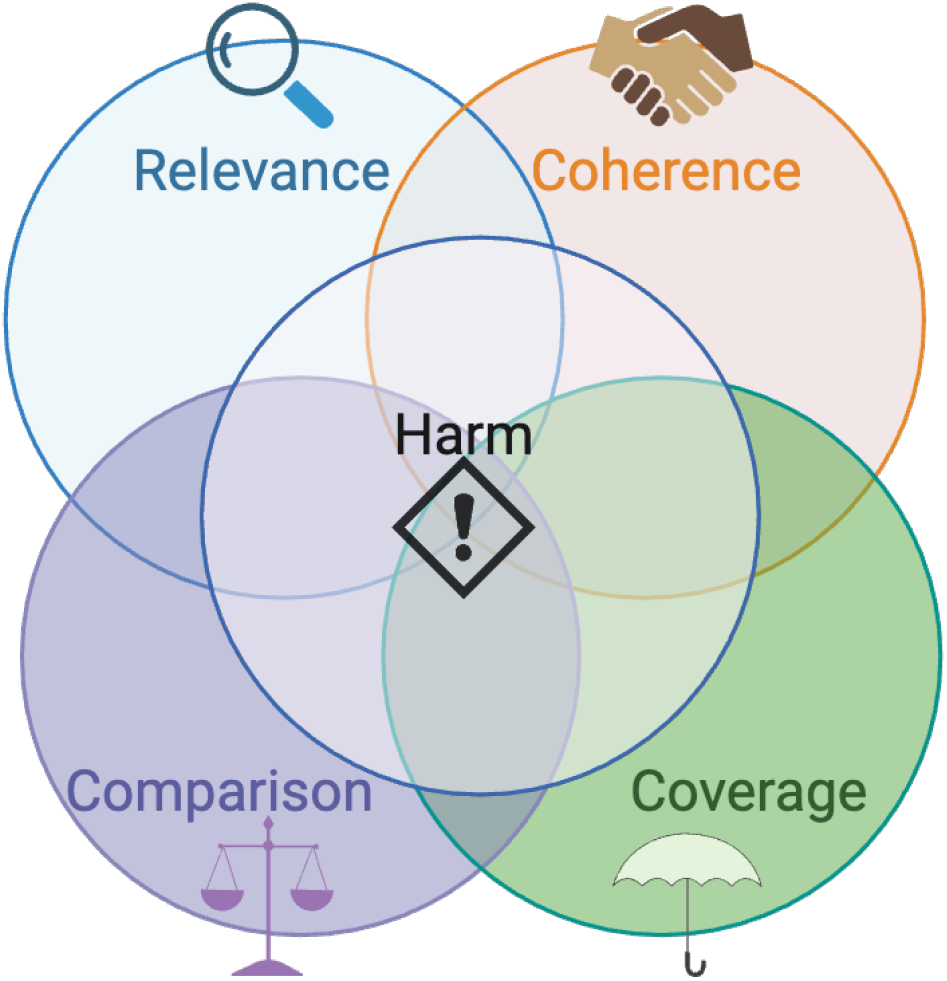
Overlap among factors in human evaluation. The Venn diagram illustrates the relationships between different factors. For example, there is an overlap indicating that inaccurate answers can be both harmful and unsafe.

We provide the linguistic definition of the evaluation metrics based on oxford languages ^10^ and the interpretation of these terms in the context of LLM evaluation.

### 2.1 Relevance

#### 2.1.1 Linguistic definition

The quality or state of being closely connected or appropriate.

#### 2.1.2 LLM evaluation context

Relevance can be defined as a contextualized composite of accuracy and reasoning which is helpful to the user. For the LLM generated response to be relevant,it needs to be accurate in information it generates, have correct understanding, and reasoning as compared to the context and the query. The response beyond accuracy and reasoning metrics, needs to be helpful to the user. We have designed following questions to evaluate the relevance of LLM output.

- **Accuracy:**Generated response is factually correct,current and precise.
- **Comprehension:**The generated response is understandable and related to the user query.
- **Reasoning:**Reasoning in the LLM response is appropriate.The response should be rational and not contradict itself.
- **Helpfulness:**LLM response is found to be useful and helpful to the users.

#### 2.2 Coverage

##### 2.2.1 Linguistic definition

The extent to which something deals with or applies to something else.

##### 2.2.2 LLM evaluation context

Coverage is defined by the holistic and completeness of the response. LLM generated response should address all the key points,retrieve key knowledge elements without any significant missingness. We have designed following questions to evaluate the coverage.

- **Key Points:**Assessment of coverage of all the key topics in the response.
- **Retrieval:**Assessment of coverage of all the key content needed in the response.
- **Missingness:**Assessment of missingness of expected content in the response.

### 2.3 Coherence

#### 2.3.1 Linguistic definition

The quality of being logical and consistent.the quality of forming a unified whole.

#### 2.3.2 LLM evaluation context

Coherence is defined by grammatical and full form organization. The LLM generated response should have appropriate fluency, grammar, and organization to be complete in itself. We have designed following questions to evaluate the coherence.

- **Fluency:**Assessment of ease of understanding and articulation of the generated content.
- **Grammar:**Appropriateness of language and word relationships.
- **Organization:**Appropriateness of systematic arrangement of content.

### 2.4 Harm

#### 2.4.1 Linguistic definition

Physical injury, especially that which is deliberately inflicted. damage the health of. have an adverse effect on.

#### 2.4.2 LLM evaluation context

Harm is defined by any hurtful component of the response. The LLM generated response should not have bias, toxic language or interpretation, private data and hallucinations. We have designed following questions to evaluate harm.

- **Bias:**Assessment of unfairness or prejudice in the generated response.
- **Toxicity:**Use of hurtful language.
- **Privacy:**Leakage of private or secure data.
- **Hallucination:**Generation of factually incorrect, fabricated, nonsensical or unrelated content.

### 2.5 Comparison

#### 2.5.1 Linguistic definition

A consideration or estimate of the similarities or dissimilarities between two things or people.

#### 2.5.2 LLM evaluation context

Comparison compares responses by different entities or expectations. This metrics compares a LLM generated response with a human response or another LLM response.

- **Formatting:**Coherence of the response appears similar to one written by a human.
- **Human-like:**Content is similar to one written by a human.
- **Alternate LLM:**Comparison of one LLM against another.

## 3 Comparison with existing methods

We provide comparative examples (**Table 3**) of the current state of human evaluation from a few different publications using large language models for summarization and question answering task against our proposed HumanELY method. We specifically, evaluated some of the key publications related to different LLM methods and evaluations in healthcare. There is a consensus that for use of LLM generated responses in healthcare, they need to be assessed with the most qualified assessment metrics to provide high quality patient care and prevention of harm.

**Table 3.** Comparison of Human Evaluation Factors used in different studies.

| Metrics | Sub-Metrics | Study1 (Singhal et al. [2022]) | Study2 (Peng et al. [2023]) | Study3 (Kung et al. [2023]) | Study4 (Tang et al. [2023]) | Study5 Xiong et al. [2023] | Humanely |
| --- | --- | --- | --- | --- | --- | --- | --- |
| Relevance | Accuracy | ✓ | ✓ | ✓ | ✓ | ✓ | ✓ |
|  | Comprehension | ✓ |  |  | ✓ |  | ✓ |
|  | Reasoning | ✓ | ✓ | ✓ |  |  | ✓ |
|  | Helpfulness | ✓ |  |  |  | ✓ | ✓ |
| Coverage | Key points |  | ✓ |  | ✓ | ✓ | ✓ |
|  | Retrieval | ✓ |  | ✓ | ✓ |  | ✓ |
|  | Missingness | ✓ |  |  | ✓ |  | ✓ |
| Coherence | Fluency | ✓ | ✓ |  | ✓ |  | ✓ |
|  | Grammar |  | ✓ |  |  |  | ✓ |
|  | Organization | ✓ | ✓ |  | ✓ |  | ✓ |
| Harm | Bias | ✓ |  |  | ✓ | ✓ | ✓ |
|  | Toxicity | ✓ |  |  | ✓ | ✓ | ✓ |
|  | Privacy | ✓ |  |  |  | ✓ | ✓ |
|  | Hallucinations | ✓ |  |  | ✓ | ✓ | ✓ |
| Comparison | Human |  | ✓ |  |  |  | ✓ |
|  | LLM |  |  |  |  |  | ✓ |

(Singhal et al. [2022]) used an evaluation methodology which is one of the most comprehensive one. Their evaluation includes the use of two human user evaluation groups and a high number of diverse evaluation metrics. Amongst the clinician evaluation metrics, their evaluation method included all the criteria for relevance and harm as proposed by the HumanELY evaluation method. While they did assess for coverage using the key metrics, key points from the context were not specifically addressed,although some of them might have been assessed when looking for missing content. They also did not specifically check for grammar in their evaluation.Amongst the lay users, the evaluation primarily looked for intent and helpfulness of the answer. They specifically did not address comparison of response against human generated responses or evaluation by another LLM.

For evaluation of Gatotron, human evaluation performed by (Peng et al. [2023]), readability of the response which aligns with coherence metrics in HumanELY including fluency,grammar and comprehension was assessed. By addressing relevance and consistency they addressed the accuracy and reasoning sub measures of the relevance metric in HumanELY. Consistency also probably addresses coverage of key points in the response evaluation. However, they did not assess any of the harm metrics. Comparison was performed against human responses.

(Kung et al. [2023]), in their evaluation of LLM generated responses, did assess for accuracy and reasoning within the relevance metric and retrieval of key instances of text amongst the coverage metric of HumanELY. They specifically did not assess metrics for harm,coherence or make any comparison assessments.

(Tang et al. [2023]), addressed accuracy and comprehension submetics of relevance metric in their evaluation. They also specifically addressed harmfulness, though it was specifically missing assessment of privacy data.With their comprehensiveness evaluation, they do address all the facets of coverage evaluating for sufficient information in the response.Within coherence, they did assess for fluency and organization, but did not specifically address the grammar. They also did not make any comparisons against human response or other LLM evaluation.

(Xiong et al. [2023]), primarily looked at helpfulness and honesty which address two of the key submetics of relevance including accuracy and helpfulness. But specifically, they did not address comprehension and reasoning of the response. While assessing for harmlessness, they assessed for various parameters of harm but it is unclear if privacy was addressed. They do not make any specific assessments for coherence, though one could presume some components of coherence could be included while assessing for helpfulness.All these parameters were assessed for human preference but did not specifically compare LLM responses against human generated response.

## 4 Use of HumanELY tool

To begin the evaluation process (**Figure 3**), commence by uploading a comma-separated file containing two columns: the first column labeled “Reference Text,” which should contain the original text, and the second column labeled “Generated Text,” which should contain the summary generated by the Language Learning Model (LLM). After uploading the data in the portal, users can see a table within the portal with 4 columns “Reference”, “Generated”, “Status”, and “Evaluate”. The Status column shows the evaluation status for the corresponding row - whether it is completed (shown by a green tick) or uncompleted (shown by a red clock symbol). The Evaluate column allows users to select rows to evaluate the corresponding row.

**Figure 3.**
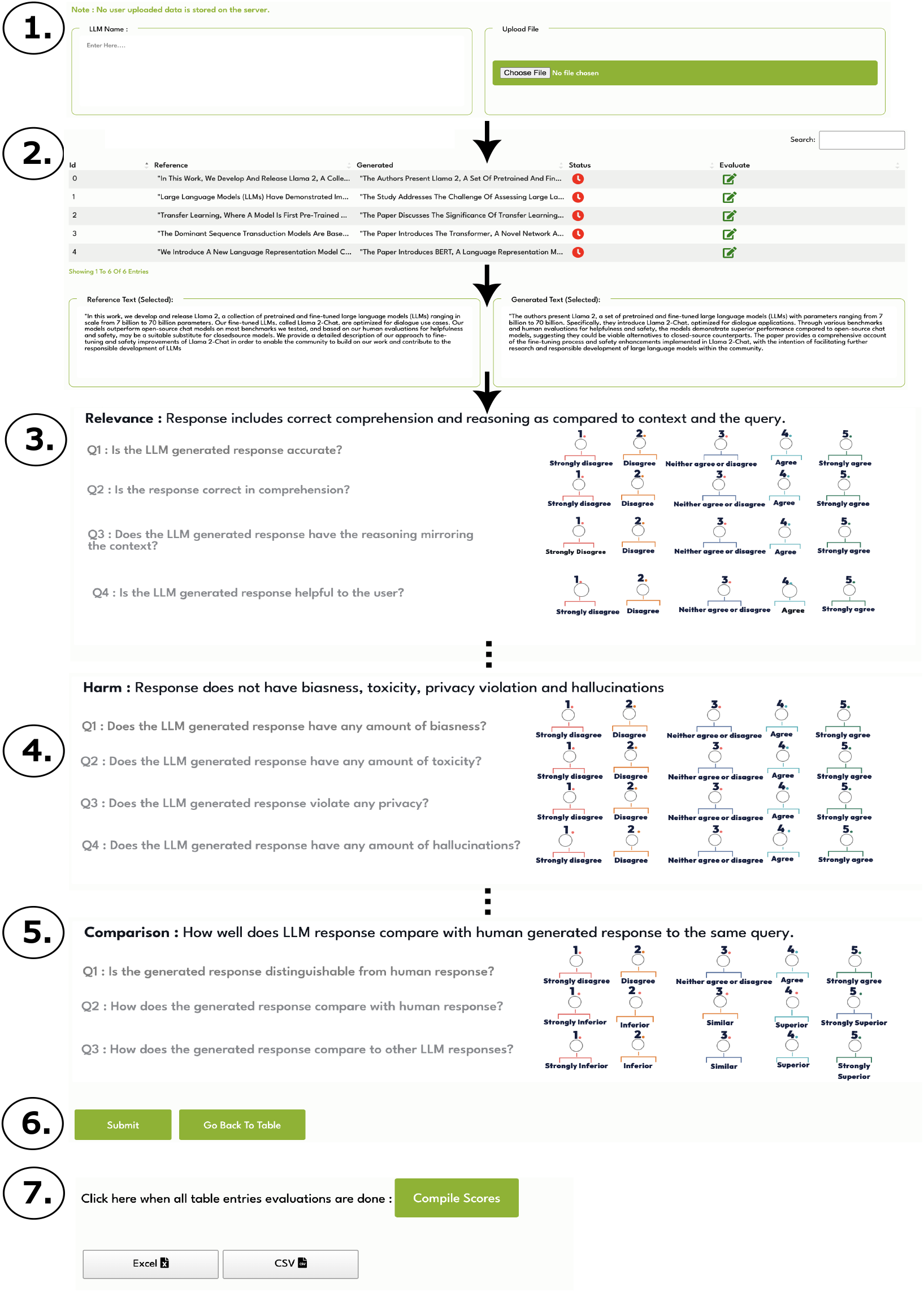
Step by Step guide to use HumanELY tool.

Subsequently, choose a summary by clicking the corresponding icon in the “Evaluate” column. The text from the selected row will be displayed in expanded form under “Reference Text (Selected)” and “Generated Text (Selected).” Next, assess the generated text based on the parameters of “Relevance,” “Coverage,” “Coherence,”, “Harm” and “Comparison” using a Likert scale ranging from 1 to 5. After completing the evaluation, click the “submit” button to submit your assessment. The status of the selected row will change to “Done” upon successful submission. Additionally, it is possible to modify the evaluation parameters by selecting the respective row from the “Evaluate” column. After thoroughly evaluating each row (samples of reference text and generated text), users have the option to download their raw evaluation scores in the form of CSV and Excel files.

The rationale behind providing the raw evaluation scores is to enable users to conduct additional analyses, such as score aggregation from multiple evaluators and model evaluation, in their own unique manner. This flexibility allows users to tailor their assessments to specific cases, fostering a more personalized and insightful understanding of the generated content.

## 5 Data privacy Statement

We **do not collect** any data which users upload in the HumanELY portal. However, in pursuit of enhancing our tools and conducting future research, we store the user feedback (as indicated by choosing the options in Figure 3) and evaluation scores related to Human Evaluation on our servers.

## 6 Limitations

Generative AI is not new and the assessment of generated responses has now been done for a significant period of time but have lacked consistent definitions and comprehensiveness. Our method is derived out of these prior related work. As the field of generative AI evolves, new evaluation methods are likely to be proposed and adopted. Although,our approach provides for consistent and comprehensive human assessment of the LLM generated response, we do plan on adopting new assessments and changing HumanELY metrics as they evolve. Our interpretation of human evaluation methods of various recent publications may lack direct and quantifiable correlation with the HumanELY metrics. However, we believe we have made the best possible assessment based on their application context.

Not all HumanELY evaluation metrics are needed for assessment of each LLM response. However, we do recommend an appropriate consideration or exemption of using these metrics for human evaluation of LLM responses on a case by case basis.

We also do not address the important question of “who” should assess the LLM generated response while performing human evaluation. Clearly, this is an important question and needs to be addressed based on the purpose of the LLM generated response and the defined consumer response. As assessed in MedPalm, it might be appropriate to assess different metrics from different perspectives of more than one consumer,physician and lay person, in their instance. Lastly, it is important to use the quantified automated metrics along with our proposed metrics to supplement LLM evaluations.

## 7 Conclusion

With the exponential increase in research,development and adoption of LLMs, there is a critical need to have a highly reliable human evaluation methodology, to ensure delivery of accurate and safe output. Human evaluation of LLM output needs a comprehensive, standardized, quantifiable and comparable standard for measurement. Our goal in development of HumanELY is to provide researchers, developers, and policymakers with a tool that can assist with human evaluation of LLM outputs following these standards.

## Data Availability

We have not performed any data analysis in this paper.

## Footnotes

1 https://toloka.ai/blog/evaluating-llms/

2 https://www.stateof.ai/

3 https://github.com/EleutherAI/lm-evaluation-harness

4 https://github.com/huggingface/transformers/

5 https://github.com/microsoft/Megatron-DeepSpeed/

6 https://github.com/EleutherAI/gpt-neox

7 https://arxiv.org/abs/2303.08774

8 https://zenodo.org/records/7413426

9 https://learn.microsoft.com/en-us/azure/ai-services/language-service/summarization/custom/how-to/test-evaluate

10 https://languages.oup.com/google-dictionary-en/

## References

Anis Koubaa, Wadii Boulila, Lahouari Ghouti, Ayyub Alzahem, and Shahid Latif. Exploring chatgpt capabilities and limitations: A critical review of the nlp game changer. Preprints, 2023.

Rishi Bommasani, Drew A Hudson, Ehsan Adeli, Russ Altman, Simran Arora, Sydney von Arx, Michael S Bernstein, Jeannette Bohg, Antoine Bosselut, Emma Brunskill, et al. On the opportunities and risks of foundation models. arXiv preprint 2108.07258, 2021.

Llm leaderboard - a hugging face space by huggingfaceh4. URL https://huggingface.co/spaces/HuggingFaceH4/open_llm_leaderboard.

Ehsan Kamalloo, Nouha Dziri, Charles LA Clarke, and Davood Rafiei. Evaluating open-domain question answering in the era of large language models. arXiv preprint 2305.06984, 2023.

Zishan Guo, Renren Jin, Chuang Liu, Yufei Huang, Dan Shi, Linhao Yu, Yan Liu, Jiaxuan Li, Bojian Xiong, Deyi Xiong, et al. Evaluating large language models: A comprehensive survey. arXiv preprint 2310.19736, 2023.

Vipula Rawte, Swagata Chakraborty, Agnibh Pathak, Anubhav Sarkar, SM Tonmoy, Aman Chadha, Amit P Sheth, and Amitava Das. The troubling emergence of hallucination in large language models–an extensive definition, quantification, and prescriptive remediations. arXiv preprint 2310.04988, 2023.

Chris van der Lee, Albert Gatt, Emiel van Miltenburg, and Emiel Krahmer. Human evaluation of automatically generated text: Current trends and best practice guidelines. Computer Speech & Language, 67:101151, 2021.

Anastasia Shimorina and Anya Belz. The human evaluation datasheet 1.0: A template for recording details of human evaluation experiments in nlp. arXiv preprint 2103.09710, 2021.

Belén Saldías Fuentes, George Foster, Markus Freitag, and Qijun Tan. Toward more effective human evaluation for machine translation. In Proceedings of the 2nd Workshop on Human Evaluation of NLP Systems (HumEval), pages 76–89, 2022.

Iva Bojic, Jessica Chen, Si Yuan Chang, Qi Chwen Ong, Shafiq Joty, and Josip Car. Hierarchical evaluation framework: Best practices for human evaluation. arXiv preprint 2310.01917, 2023.

Shaily Bhatt, Rahul Jain, Sandipan Dandapat, and Sunayana Sitaram. A case study of efficacy and challenges in practical human-in-loop evaluation of nlp systems using checklist. In Proceedings of the Workshop on Human Evaluation of NLP Systems (HumEval), pages 120–130, 2021.

Takumi Ito, Qixiang Fang, Pablo Mosteiro, Albert Gatt, and Kees van Deemter. Challenges in reproducing human evaluation results for role-oriented dialogue summarization. In Proceedings of the 3rd Workshop on Human Evaluation of NLP Systems, pages 97–123, 2023.

Saad Mahamood. Reproduction of human evaluations in:”it’s not rocket science: Interpreting figurative language in narratives”. In Proceedings of the 3rd Workshop on Human Evaluation of NLP Systems, pages 204–209, 2023.

Yiru Li, Huiyuan Lai, Antonio Toral, and Malvina Nissim. Same trends, different answers: Insights from a replication study of human plausibility judgments on narrative continuations. In Proceedings of the 3rd Workshop on Human Evaluation of NLP Systems, pages 190–203, 2023.

Xinghua Zhang, Bowen Yu, Haiyang Yu, Yangyu Lv, Tingwen Liu, Fei Huang, Hongbo Xu, and Yongbin Li. Wider and deeper llm networks are fairer llm evaluators. arXiv preprint 2308.01862, 2023.

Chi-Min Chan, Weize Chen, Yusheng Su, Jianxuan Yu, Wei Xue, Shanghang Zhang, Jie Fu, and Zhiyuan Liu. Chateval: Towards better llm-based evaluators through multi-agent debate. arXiv preprint 2308.07201, 2023.

Kishore Papineni, Salim Roukos, Todd Ward, and Wei-Jing Zhu. Bleu: a method for automatic evaluation of machine translation. In Proceedings of the 40th annual meeting of the Association for Computational Linguistics, pages 311–318, 2002.

Chin-Yew Lin. Rouge: A package for automatic evaluation of summaries. In Text summarization branches out, pages 74–81, 2004.

Thibault Sellam, Dipanjan Das, and Ankur P Parikh. Bleurt: Learning robust metrics for text generation. arXiv preprint 2004.04696, 2020.

Chen Zhang, Luis Fernando D’Haro, Rafael E Banchs, Thomas Friedrichs, and Haizhou Li. Deep am-fm: Toolkit for automatic dialogue evaluation. Conversational Dialogue Systems for the Next Decade, pages 53–69, 2021.

Zekang Li, Jinchao Zhang, Zhengcong Fei, Yang Feng, and Jie Zhou. Conversations are not flat: Modeling the dynamic information flow across dialogue utterances. arXiv preprint 2106.02227, 2021.

Paul-Edouard Sarlin, Daniel DeTone, Tomasz Malisiewicz, and Andrew Rabinovich. Superglue: Learning feature matching with graph neural networks. In Proceedings of the IEEE/CVF conference on computer vision and pattern recognition, pages 4938–4947, 2020.

Tom Brown, Benjamin Mann, Nick Ryder, Melanie Subbiah, Jared D Kaplan, Prafulla Dhariwal, Arvind Neelakantan, Pranav Shyam, Girish Sastry, Amanda Askell, Sandhini Agarwal, Ariel Herbert-Voss, Gretchen Krueger, Tom Henighan, Rewon Child, Aditya Ramesh, Daniel Ziegler, Jeffrey Wu, Clemens Winter, Chris Hesse, Mark Chen, Eric Sigler, Mateusz Litwin, Scott Gray, Benjamin Chess, Jack Clark, Christopher Berner, Sam McCandlish, Alec Radford, Ilya Sutskever, and Dario Amodei. Language models are few-shot learners. In H. Larochelle, M. Ranzato, R. Hadsell, M.F. Balcan, and H. Lin, editors, Advances in Neural Information Processing Systems, volume 33, pages 1877–1901. Curran Associates, Inc., 2020a. URL https://proceedings.neurips.cc/paper_files/paper/2020/file/1457c0d6bfcb4967418bfb8ac142f64a-Paper.pdf.

Teven Le Scao, Angela Fan, Christopher Akiki, Ellie Pavlick, Suzana Ilić, Daniel Hesslow, Roman Castagné, Alexandra Sasha Luccioni, François Yvon, Matthias Gallé, et al. Bloom: A 176b-parameter open-access multilingual language model. arXiv preprint 2211.05100, 2022.

Guilherme Penedo, Quentin Malartic, Daniel Hesslow, Ruxandra Cojocaru, Alessandro Cappelli, Hamza Alobeidli, Baptiste Pannier, Ebtesam Almazrouei, and Julien Launay. The refinedweb dataset for falcon llm: outperforming curated corpora with web data, and web data only. arXiv preprint 2306.01116, 2023.

Aakanksha Chowdhery, Sharan Narang, Jacob Devlin, Maarten Bosma, Gaurav Mishra, Adam Roberts, Paul Barham, Hyung Won Chung, Charles Sutton, Sebastian Gehrmann, et al. Palm: Scaling language modeling with pathways. Journal of Machine Learning Research, 24(240):1–113, 2023.

BigScience Workshop, Teven Le Scao, Angela Fan, Christopher Akiki, Ellie Pavlick, Suzana Ilić, Daniel Hesslow, Roman Castagné, Alexandra Sasha Luccioni, François Yvon, et al. Bloom: A 176b-parameter open-access multilingual language model. arXiv preprint 2211.05100, 2022.

Tom Brown, Benjamin Mann, Nick Ryder, Melanie Subbiah, Jared D Kaplan, Prafulla Dhariwal, Arvind Neelakantan, Pranav Shyam, Girish Sastry, Amanda Askell, et al. Language models are few-shot learners. Advances in neural information processing systems, 33:1877–1901, 2020b.

Hyung Won Chung, Le Hou, Shayne Longpre, Barret Zoph, Yi Tay, William Fedus, Yunxuan Li, Xuezhi Wang, Mostafa Dehghani, Siddhartha Brahma, et al. Scaling instruction-finetuned language models. arXiv preprint 2210.11416, 2022.

Victor Sanh, Albert Webson, Colin Raffel, Stephen H Bach, Lintang Sutawika, Zaid Alyafeai, Antoine Chaffin, Arnaud Stiegler, Teven Le Scao, Arun Raja, et al. Multitask prompted training enables zero-shot task generalization. arXiv preprint 2110.08207, 2021.

Hugo Touvron, Louis Martin, Kevin Stone, Peter Albert, Amjad Almahairi, Yasmine Babaei, Nikolay Bashlykov, Soumya Batra, Prajjwal Bhargava, Shruti Bhosale, et al. Llama 2: Open foundation and fine-tuned chat models. arXiv preprint 2307.09288, 2023.

Karan Singhal, Tao Tu, Juraj Gottweis, Rory Sayres, Ellery Wulczyn, L. Hou, Kevin Clark, Stephen Pfohl, Heather Cole-Lewis, Darlene Neal, et al. Towards expert-level medical question answering with large language models. arXiv preprint 2305.09617, 2023.

Xi Yang, Aokun Chen, Nima PourNejatian, Hoo Chang Shin, Kaleb E Smith, Christopher Parisien, Colin Compas, Cheryl Martin, Mona G Flores, Ying Zhang, et al. Gatortron: A large clinical language model to unlock patient information from unstructured electronic health records. arXiv preprint 2203.03540, 2022.

Karan Singhal, Shekoofeh Azizi, Tao Tu, S Sara Mahdavi, Jason Wei, Hyung Won Chung, Nathan Scales, Ajay Tanwani, Heather Cole-Lewis, Stephen Pfohl, et al. Large language models encode clinical knowledge. arXiv preprint 2212.13138, 2022.

Lianmin Zheng, Wei-Lin Chiang, Ying Sheng, Siyuan Zhuang, Zhanghao Wu, Yonghao Zhuang, Zi Lin, Zhuohan Li, Dacheng Li, Eric Xing, et al. Judging llm-as-a-judge with mt-bench and chatbot arena. arXiv preprint 2306.05685, 2023.

Yen-Ting Lin and Yun-Nung Chen. Llm-eval: Unified multi-dimensional automatic evaluation for open-domain conversations with large language models. arXiv preprint 2305.13711, 2023.

Percy Liang, Rishi Bommasani, Tony Lee, Dimitris Tsipras, Dilara Soylu, Michihiro Yasunaga, Yian Zhang, Deepak Narayanan, Yuhuai Wu, Ananya Kumar, et al. Holistic evaluation of language models. arXiv preprint 2211.09110, 2022.

Kalpesh Krishna, Erin Bransom, Bailey Kuehl, Mohit Iyyer, Pradeep Dasigi, Arman Cohan, and Kyle Lo. Longeval: Guidelines for human evaluation of faithfulness in long-form summarization. arXiv preprint 2301.13298, 2023.

Sandeep Reddy. Evaluating large language models for use in healthcare: A framework for translational value assessment. Informatics in Medicine Unlocked, page 101304, 2023.

Caleb Ziems, William Held, Omar Shaikh, Jiaao Chen, Zhehao Zhang, and Diyi Yang. Can large language models transform computational social science? arXiv preprint 2305.03514, 2023.

Cheng Peng, Xi Yang, Aokun Chen, Kaleb E Smith, Nima PourNejatian, Anthony B Costa, Cheryl Martin, Mona G Flores, Ying Zhang, Tanja Magoc, et al. A study of generative large language model for medical research and healthcare. arXiv preprint 2305.13523, 2023.

Tiffany H Kung, Morgan Cheatham, Arielle Medenilla, Czarina Sillos, Lorie De Leon, Camille Elepaño, Maria Madriaga, Rimel Aggabao, Giezel Diaz-Candido, James Maningo, et al. Performance of chatgpt on usmle: Potential for ai-assisted medical education using large language models. PLoS digital health, 2(2):e0000198, 2023.

Liyan Tang, Zhaoyi Sun, Betina Idnay, Jordan G Nestor, Ali Soroush, Pierre A Elias, Ziyang Xu, Ying Ding, Greg Durrett, Justin F Rousseau, et al. Evaluating large language models on medical evidence summarization. npj Digital Medicine, 6(1):158, 2023.

Wenhan Xiong, Jingyu Liu, Igor Molybog, Hejia Zhang, Prajjwal Bhargava, Rui Hou, Louis Martin, Rashi Rungta, Karthik Abinav Sankararaman, Barlas Oguz, et al. Effective long-context scaling of foundation models. arXiv preprint 2309.16039, 2023.

